# Evaluation of the ID NOW Among Symptomatic Individuals During the Omicron Wave

**DOI:** 10.1101/2022.05.19.22275316

**Authors:** William Stokes, Allison A. Venner, Emily Buss, Graham Tipples, Byron M. Berenger

## Abstract

**BACKGROUND:** Point of Care SARS-CoV-2 devices, such as the Abbott ID NOW have great potential, to help combat the COVID-19 pandemic. Starting in December, 2020, the ID NOW was implemented throughout the province of Alberta, Canada (population 4.4 million) in various settings. We aimed to assess the ID NOW performance during the BA.1 Omicron wave and compare it to previous waves.

**METHODS:** The ID NOW was assessed in two distinct locations among symptomatic individuals: acute care (emergency room, urgent care, and hospitalized patients) and community assessment centres (AC) during the period January 5 – 18, 2022. Starting January 5, Omicron represented >95% of variants detected in our population. For every individual tested, two swabs were collected: one for ID NOW testing and the other for either reverse-transcriptase polymerase chain reaction (RT-PCR) confirmation of negative ID NOW results or for variant testing of positive ID NOW results.

**RESULTS:** A total of 3,041 paired samples were analyzed (1,139 RT-PCR positive). 1,873 samples were from 42 COVID-19 AC and 1,168 from 69 rural hospitals. ID NOW sensitivity for symptomatic individuals presenting to community AC and patients in hospital was 96.0% [95% confidence interval (CI) 94.5-97.3%, n=830 RT-PCR positive], and 91.6% (95% CI 87.9-94.4%, n=309 RT-PCR positive), respectively. SARS-CoV 2 positivity rate was very high for both populations (44.3% at AC, 26.5% in hospital).

**CONCLUSIONS:** Sensitivity of ID NOW SARS-CoV-2, compared to RT-PCR, is very high during the BA.1 Omicron wave, and is significantly higher when compared to previous SARS-CoV-2 variant waves.

## INTRODUCTION

The ID NOW (Abbott, Illinois, United States) is approved by the United States Food and Drug Administration (FDA) Emergency Use Authorization for the point of care, rapid detection of severe acute respiratory syndrome-coronavirus-2 (SARS-CoV-2) in individuals who are within the first 7 days of symptom onset.^1^ The ID NOW assay uses isothermal nucleic acid amplification of a region of the viral RNA-dependent RNA polymerase (RdRp) to detect the presence of SARS-CoV-2, with results available in under 15 minutes. Clinical specimens approved by U.S. FDA for testing include nasal, oropharyngeal (OP), and nasopharyngeal swabs (NP), which must be tested on the Abbott ID NOW either immediately or within one hour of collection. Specimens placed in viral/universal transport media (UTM) are not valid for testing by the Abbott ID NOW.^1^

Since December 4, 2020 the ID NOW has been used in various settings throughout Alberta, Canada (4.4 million people). This includes 42 COVID-19 assessment centres and 69 rural hospitals. We previously prospectively evaluated the ID NOW performance among symptomatic individuals in these sites prior to the Omicron variant wave. The clinical sensitivity and specificity of the ID NOW among symptomatic individuals presenting to assessment centres (community swab centres) was 92.5% [95% confidence interval (CI) 92.0-93.0%] and 99.5% (95% CI 99.4-99.5%), respectively (n=70,879 with 10,633 RT-PCR positive). The clinical sensitivity and specificity of the ID NOW among symptomatic individuals in hospital was 89.5% (95% CI 88.3-90.6%) and 99.3% (95% CI 99.2-99.4%), respectively (n=16,924 with 2,932 RT-PCR positive).^2^

Currently, there is a paucity of data on the ID NOW performance in the setting of the Omicron variant. Abbott has reported that mutations found in the Omicron variant does not affect the performance based on *in silico* data.^3^ Due to the different growth kinetics, transmissibility and tissue tropisms than other variants and emergence in a population with 72% vaccination^4^, the performance with Omicron may be different and clinical validation is justified.^5^

Our aim was to assess ID NOW’s clinical performance among symptomatic individuals presenting to assessment centres and patients in emergency rooms and hospitals during a time when the Omicron variant represented the majority of SARS-CoV-2 detected within our population.

## METHODS

Since December 4, 2020, the ID NOW was gradually implemented across Alberta in the following sites:

1. 42 COVID-19 Alberta Health Services (AHS) Public Health assessment/swabbing centres, located in all regions of Alberta: testing of symptomatic individuals and asymptomatic close contacts. These are the primary locations for community patients not needing medical attention to get tested for COVID-19 in Alberta. Testing and swabbing was performed by assessment centre staff (e.g. nurses) within Alberta Precision Laboratories (APL) approved Point of Care Testing (POCT) programs.
2. 69 rural hospitals located across Alberta: testing of symptomatic inpatients or ED patients. Swabbing was performed by physicians, nurses, or respiratory therapists. Testing was performed in a College of Physician and Surgeons of Alberta (CPSA) accredited hospital laboratory (APL). The collection and testing of COVID-19 samples followed the same procedures as described in our previous paper.^2^ We chose to analyze data from January 5 – 18, 2022, when Omicron represented >95% of SARS-CoV-2 variants detected in our population. Not all individuals with ID NOW positive samples had samples collected for testing of variants of concern, due to changes in our testing protocols designed to conserved lab capacity.

Individuals were given the option to have POCT by ID NOW and routine testing, or routine testing alone. All individuals tested with the ID NOW had two parallel swabs collected. The first swab collected was either a NP swab or OP swab, which was placed in UTM (Yocon Biology, Beijing, China or GDL Korea Co. Ltd, Seoul, Korea) for RT-PCR, and transported to an accredited laboratory at room temperature and stored at 4°C until processing. The second swab was an OP swab for ID NOW testing (using the OP swab provided in the ID NOW kits). The OP swab for ID NOW testing was always collected second to ensure all individuals had a sample available for RT-PCR (i.e. in case the individual refused the second NP or OP swab). If the ID NOW test was negative, the second swab was sent for confirmatory RT-PCR testing. If the ID NOW test was positive, the second swab was either sent for storage or for variant of concern screening (VoC) testing. All RT-PCR tests sent for variant testing were done within approximately 72 hours from time of collection. During the Omicron wave, only a subset of positive samples were sent for variant testing due to resourcing pressures faced by our laboratory. Positive ID NOW results without subsequent variant testing were considered true positives in this study.

All RT-PCR testing was performed on the APL Public Health Laboratory E gene PCR or on a Health Canada and FDA approved commercial assay.^6^ Commercial assays were dependent on the testing lab and included the Allplex (Seegene, Seoul, South Korea), BDMax (Becton Dickinson, NJ, United States), Panther (Hologic, MA, United States), GeneXpert SARS-CoV-2 or SARS-CoV-2/Influenza/RSV (Cepheid, CA, United States), Cobas 6800 System (Roche Molecular Systems, CA, United States) and Simplexa (DiaSorin, Saluggia, Italy). All samples sent for VoC screening were tested with the ProvLab E gene assay to determine if sufficient viral load was present for VoC testing. E gene RT-PCR results from our lab-developed test were considered positive for SARS-CoV-2 when E gene cycle threshold (Ct) value was <35. If the Ct was ≥35, amplification from the same eluate was repeated in duplicate and was considered positive if at least 2/3 results had a Ct <41.

All personnel performing the ID NOW swabbing/testing were trained healthcare workers (HCW), who were previously trained in NP and oropharyngeal swab collection. At time of collection, they asked and recorded whether the patient had symptoms or was asymptomatic. All sites and HCW were trained on the ID NOW collection, transport, and testing processes, at least according to the manufacturer’s instructions, prior to ID NOW implementation. AHS staff were trained according to the APL POCT program, which meets CPSA accreditation standards. Each ID NOW device underwent a verification process, which included testing 3 positive ID NOW control swabs and 5 negative ID NOW control swabs on the ID NOW instrument before use. One positive control and one negative control swab were tested on the ID NOW instrument after each new box of ID NOW kits was opened, after each new HCW was trained on the instrument, and, after each instrument was transported to a different site.

Samples were included for all ID NOW results collected at assessment centres or hospitals among symptomatic individuals. Results without proper documentation of testing location, or without confirmatory RT-PCR, if ID NOW negative, were excluded.

Data was pulled from our provincial laboratory’s centralized electronic database containing SARS-CoV-2 results for all publicly funded testing in the province except for border testing. Sensitivity and specificity of the ID NOW was calculated with Clopper-Pearson 95% confidence intervals. Statistical analysis was performed using Pearson Chi-squared for categorical variables and t-test for continuous variables using STATA (version 14.1).

The University of Alberta Research Ethics board approved this study (Pro00111835).

## RESULTS

A total of 3,498 results were identified between January 5-18, 2022. 457 samples were excluded: 35 did not have testing location recorded, 24 did not have an ID NOW result recorded and 398 ID NOW negative results did not have parallel RT-PCR results recorded. The remaining 3,041 paired samples were analyzed.

Of the 3,041 paired samples, 1,873 were collected from 42 assessment centres and 1,168 from 69 rural hospitals. Baseline characteristics and variant test results of these samples are provided in Table 1. Results, compared to RT-PCR, are provided in Table 2 and Figure 1. In both assessment centres and hospitals, ID NOW positivity rate was extremely high at 44.3% and 26.5%, respectively, and is consistent with SARS-CoV-2 positivity rates observed among other COVID-19 diagnostic platforms during the same time [AHS Surveillance Data]. ID NOW sensitivity was higher among symptomatic individuals presenting to assessment centres (96.0%, 95% CI 94.5-97.3%, n=830 RT-PCR positive) compared to symptomatic patients in hospital (91.6% (95% CI 87.9-94.4%, n=309 RT-PCR positive). ID NOW specificity, negative predictive value (NPV), and positive predictive value (PPV) among symptomatic individuals presenting to assessment centres was 97.8% (95% CI 96.7-98.6%), 96.9% (95% CI 95.7-97.7%), and 97.2% (95.9-98.1%), respectively. ID NOW specificity, NPV, and PPV among symptomatic individuals in hospital was 98.6% (95% CI 97.6-99.3%), 97.0% (95.8-97.9%), and 95.9% (93.1-97.6%), respectively.

**Table 1:** Characteristics between individuals tested with ID NOW SARS-CoV-2

| Site | Assessment centre | Hospital |
| --- | --- | --- |
| Mean age (median, range) | 38.6 (38.0, 0.5-85.8) | 51.3 (55.2, 0.09-103.2) |
| Male gender | 33.1% | 48.8% |
| NP swab used for parallel RT-PCR testing* (%) | 78.3% | 92.1% |
| Variant** (%) | Omicron (89.3%)<br>Delta (0.7%)<br>Unresolved (10.0%) | Omicron (70.4%)<br>Delta (14.5%)<br>Unresolved (15.0%) |
| Mean E gene Ct value from variant testing of true positives (median, range) | 22.7 (21.6, 14.5-40.9) | 22.6 (21.5,15.2-38.1) |
| Mean E gene Ct value from variant testing of false negatives (median, range) | 25.8 (25.4, 21.6-34.5) | 32.1 (34.1, 17.1-36.7) |

NP: nasopharyngeal
Ct: Cycle threshold
\*Either NP or oropharyngeal swab was used for parallel RT-PCR testing. In hospitalized patients, a minority of samples were other specimen types such as endotracheal tube aspirates.
\*\*n=428 for assessment centres, n=193 for hospital

**Table 2:**
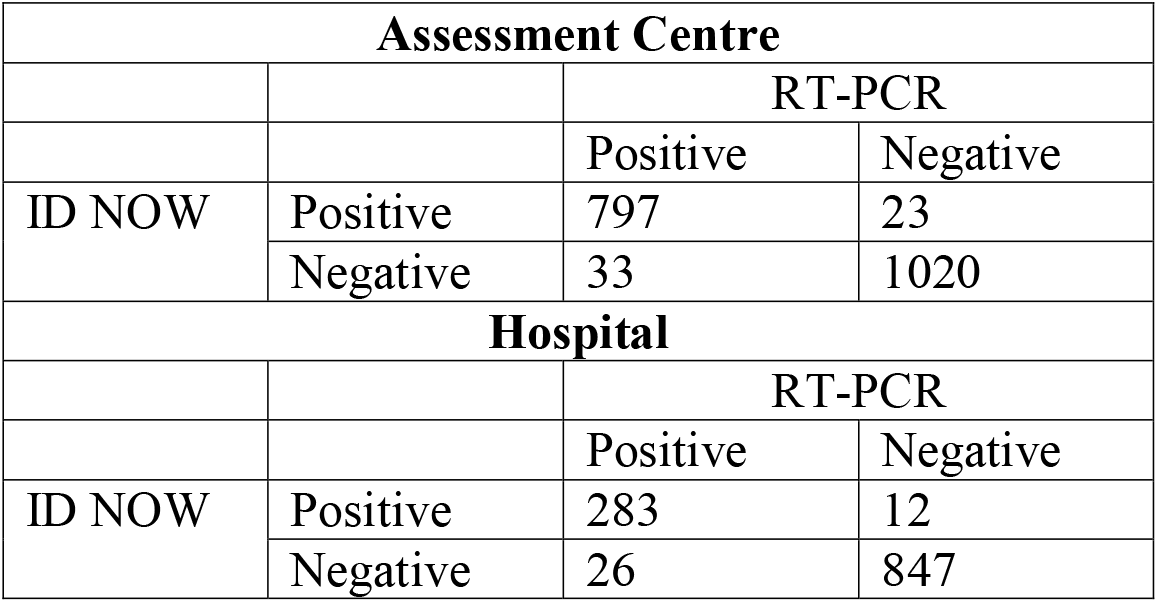
Performance of ID NOW, compared to RT-PCR, using oropharyngeal swabs.

| Assessment Centre |  |  |  |
| --- | --- | --- | --- |
|  |  | RT-PCR |  |
|  |  | Positive | Negative |
| ID NOW | Positive | 797 | 23 |
|  | Negative | 33 | 1020 |
| Hospital |  |  |  |
|  |  | RT-PCR |  |
|  |  | Positive | Negative |
| ID NOW | Positive | 283 | 12 |
|  | Negative | 26 | 847 |

**Figure 1:**
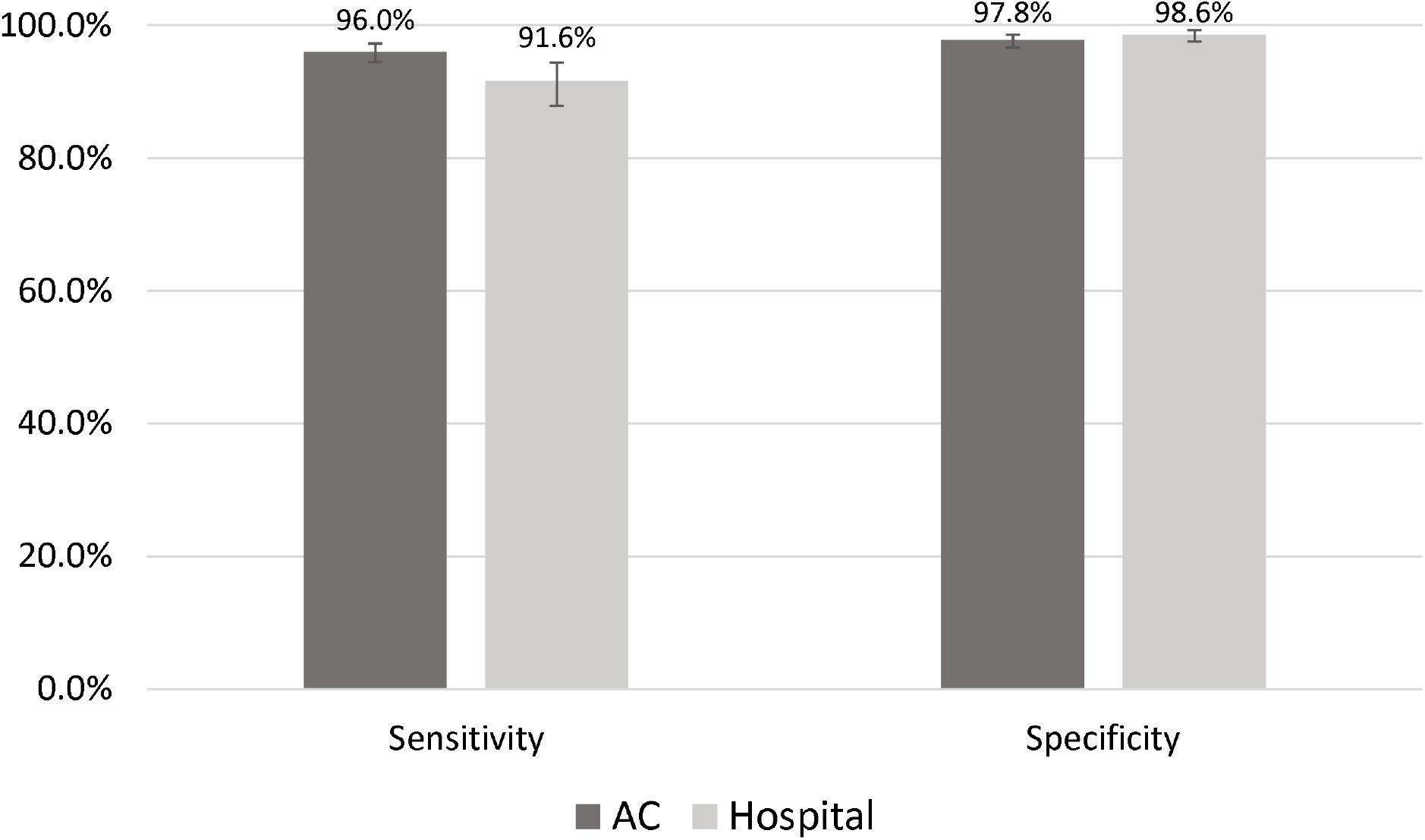
Sensitivity and specificity of ID NOW (oropharyngeal swab) compared to RT-PCR (oropharyngeal swab or nasopharyngeal swab). Error bars represent 95% confidence intervals. AC = assessment centre.

E gene Ct values for ID NOW true positive and true negative results are provided in Table 1. Mean E gene Ct values were higher among ID NOW false negatives compared to true positives, but were only statistically significant for hospital patients (p<0.001).

## DISCUSSION

Use of point of care SARS-CoV-2 tests, such as the ID NOW, for the detection of SARS-CoV-2 among individuals remains a worthwhile endeavour. At time of writing, this is the first study to evaluate the ID NOW during the Omicron variant wave. In the face of Omicron, our study demonstrated increased ID NOW sensitivity compared to prior variant waves.^2^ The ID NOW sensitivity in assessment centres before Omicron (as of November 24, 2021) was 92.5% (95% CI 92.0-93.0%). Our current data during the Omicron wave has demonstrated a higher ID NOW sensitivity of 96.0% (95% CI 94.5-97.3%). Sensitivity in hospital settings also increased with the Omicron wave, though the difference was not statistically significant (pre-Omicron it was 89.5%, 95% CI 88.3-90.6%, compared to 91.6% during Omicron, 95% CI 87.9-94.4%). In respose to this study and the high sensitivity in previous waves, our provincial laboratory no longer requires confirmation of negative ID NOW results when tested on symptomatic individuals presenting to community assessment centres within 7 days of symptom onset.

In the current Omicron study and our previous ID NOW study, sensitivity was slightly lower in hospital settings compared to community assessment centres. However, its performance was still higher for Omicron than what we previously observed, but not statistically significant (likely due to the lower sample size in our Omicron study).^2^ There may be various factors to account for the decreased ID NOW sensitivity observed among hospitals compared to assessment centres. Firstly, ID NOW testing is performed immediately after sample collection at assessment centres, whereas hospitals require transportation to the on-site lab prior to ID NOW testing. While ID NOW testing was mandated to be done within 1 hour from collection, short periods of time from transportation may potentially affect performance.^7^ Secondly, a higher proportion of individuals with lower viral loads may be tested in emergency rooms, as a result of patients commonly presenting to hospital later in their symptom onset course, which is associated with higher SARS-CoV-2 E gene Ct values.^8,9^ Symptomatic individuals presenting to assessment centres, in comparison, are often within the first few days of symptom onset, but because it generally takes approximately 24 hours to arrange a booking, are not at the very beginning of their symptom onset.

Why ID NOW sensitivity improved in the face of Omicron is not fully known. There have been reports that the SARS-CoV-2 viral loads observed among clinical specimens with the Omicron variant are not different from clinical specimens of other variants.^10,11^ There are suggestions that Omicron has altered tissue tropism compared to prior variants which may make sampling from the oropharynx more reliable.^12^ Higher COVID-19 prevalence during the Omicron wave could have also contributed. Positivity rate for all COVID-19 tests done during the study period in Alberta was 36.74% compared to 4.27% and 9.09% during the wild type peaks (April 2020 and December 2020), 13.56 for alpha (May 2021), and 13.47% for Delta (September 2021) [AHS surveillance dashboard].

Interestingly, a high proportion (87.0%) of the false positives detected in our study, among individuals at assessment centres, were from samples collected using a nasopharyngeal swab. Due to the many (45.5%) ID NOW positive samples that were subsequently not tested for variants in our study, the specificity calculated in our study is likely even lower. The reasons behind reduced specificity is unclear, but potentially related to altered tissue tropism favouring samples from the oropharynx over the nasopharynx when tested earlier in the COVID-19 infection course.13 Lower specificity is unlikely to be explained by other factors, such as SARS-CoV-2 contamination, given that our testing was conducted across over 100 sites and there has not been prior reports of contamination or high rates of false positive results with the ID NOW in the literature.

Strengths of this study include the large sample size studied in various real-world locations, including community COVID-19 assessment centres and hospitals. Due to the same testing sites, procedures, and methodology, the results during Omicron can be accurately compared to our results from before Omicron.^2^ Due to the routine surveillance testing of many positive ID NOW or RT-PCR samples for variants of concern, which included E gene RT-PCR from a consistent RT-PCR testing platform (APL LDT), we were able to roughly assess the specificity of the ID NOW and examine the relationship of E gene Ct values between true positive and false negative ID NOW results in the face of Omicron.

Our study had several limitations. Due to the heterogeneity of our populations tested, it is difficult to exclude confounders that may have contributed to ID NOW performance. However, we previously observed no differences in sensitivity based on common patient, collecting and testing characteristics, including age, gender, and swab type.^2^ Our current study also did not demonstrate any difference in performance with these characteristics, though the sample size was not large enough to make any concrete conclusions. Another limitation is the inability to exclusively study Omicron by itself. While the Delta variant only represented 0.7% of variants detected from the assessment centres during our study period, there was still a moderately high proportion (14.5%) of delta variant still circulating within our hospitals. This study did not assess the Omicron BA.2 sublineage, as it was not circulating in our population during the study period.

Other limitations include missing parallel RT-PCR results that could affect the sensitivity observed in our study. As previously mentioned, many (45.5%) ID NOW positive samples did not undergo variant testing which would affect our calculated specificity. ID NOW sensitivity may be slightly lower than we calculated due to the exclusion of 398 ID NOW negative samples that subsequently did not have a second sample tested with RT-PCR. Reasons behind missing parallel RT-PCR are multifactorial and include sample lost or discarded prior to testing, testing sites going against guidelines and not obtaining a second swab for RT-PCR confirmation, and patient demographic mismatches resulting in test cancellation or inability to match ID NOW and RT-PCR tests together in our electronic database.

In conclusion, the performance of the ID NOW improved in sensitivity during the Omicron BA.1 wave. Reasons for improved sensitivity may potentially be related to increased COVID-19 prevalence and/or altered tissue tropism favouring the oropharynx early in the COVID-19 infection course. More research is required to prove this claim.

## Data Availability

All data produced in the present study are available upon reasonable request to the authors

## Acknowledgments

This work was funded using internal operating funds of Alberta Precision Laboratories and Alberta Health Services (AHS). Test kits and instruments were paid for by the Public Health Agency of Canada. We thank staff at AHS assessment centre and contracted mobile teams for collecting and testing samples in the community, and Alberta Precision Laboratory staff for assistance with testing of samples, and in the development and support of the various testing programs.

## Declarations

### Availability of data

Available upon request.

